# A Global Perspective on Determinants of Cardiovascular Mortality: Linear Regression Model of 183 Countries on Nationwide Cardiovascular Policies, Socioeconomic Disparities, Universal Health Coverage and Tobacco

**DOI:** 10.1101/2025.04.23.25326323

**Authors:** Hamzah Pratama Megantara, Iwan Dakota, Taofan, Suci Indriani, Ruth Grace Aurora, Suko Adiarto

## Abstract

**Background:** Cardiovascular (CV) diseases remain the leading cause of mortality worldwide. Various determinants has been indicated contributing in CV mortality, spanning from health policy, universal health coverage (UHC), tobacco and socioeconomic diversity. To date, no quantitative regression model has been established to exhibit the magnitude and prediction of CV mortality. We aimed to formulate a linear regression model of CV determinants involving healthcare insurance coverage and national policy, economic status, smoking prevalence, gender and subregional variety.

**Methods:** A linear regression model was employed, with the percentage of cardiovascular deaths as the dependent variable. Independent variables were Universal Health Coverage (UHC), World Bank income classifications, availability of national policy on CV, gender, availability of national CV guideline, CV risk stratification in primary healthcare, smoking and sub-regions. Data were gathered from World Health Organization and The World Bank dataset.

**Results:** A total of 2,385 data points from 2015 to 2019 was acquired constituting 183 countries. More extensive UHC (β = -0.052, t = -3.663, p <0.001) and high-income countries (-0.060, t = -2.756, p <0.001) exhibited lower predicted CV mortality. Smoking prevalence was strongly correlated with higher mortality (β = 0.128, t = 8.408, p <0.001). Regional disparities were observed, with Eastern Europe presenting highest mortality rate (β = 0.685, t = 32.686, p <0.001). Compared to male, female showed higher cardiovascular death (β = 0.047, t = 4.017, p <0.001). The availability of national policies in cardiovascular health were associated with lower mortality (β = -0.031, t = -3.211, p = 0.001). CV national guideline was the only non-significant CV determinant.

**Conclusions:** The development of a quantitative regression model for cardiovascular mortality incorporating multifaceted determinants was expected to promote comprehensive public health strategies, policy reforms, and national health system strengthening, which are essential to reduce the global burden of cardiovascular diseases.

Figure 1
(Graphical Abstract).
Summary of the research and quantified determinants of CV death. UHC: universal health coverage, CV: cardiovascular, *≥50% availability of CV risk stratification program in primary health care.

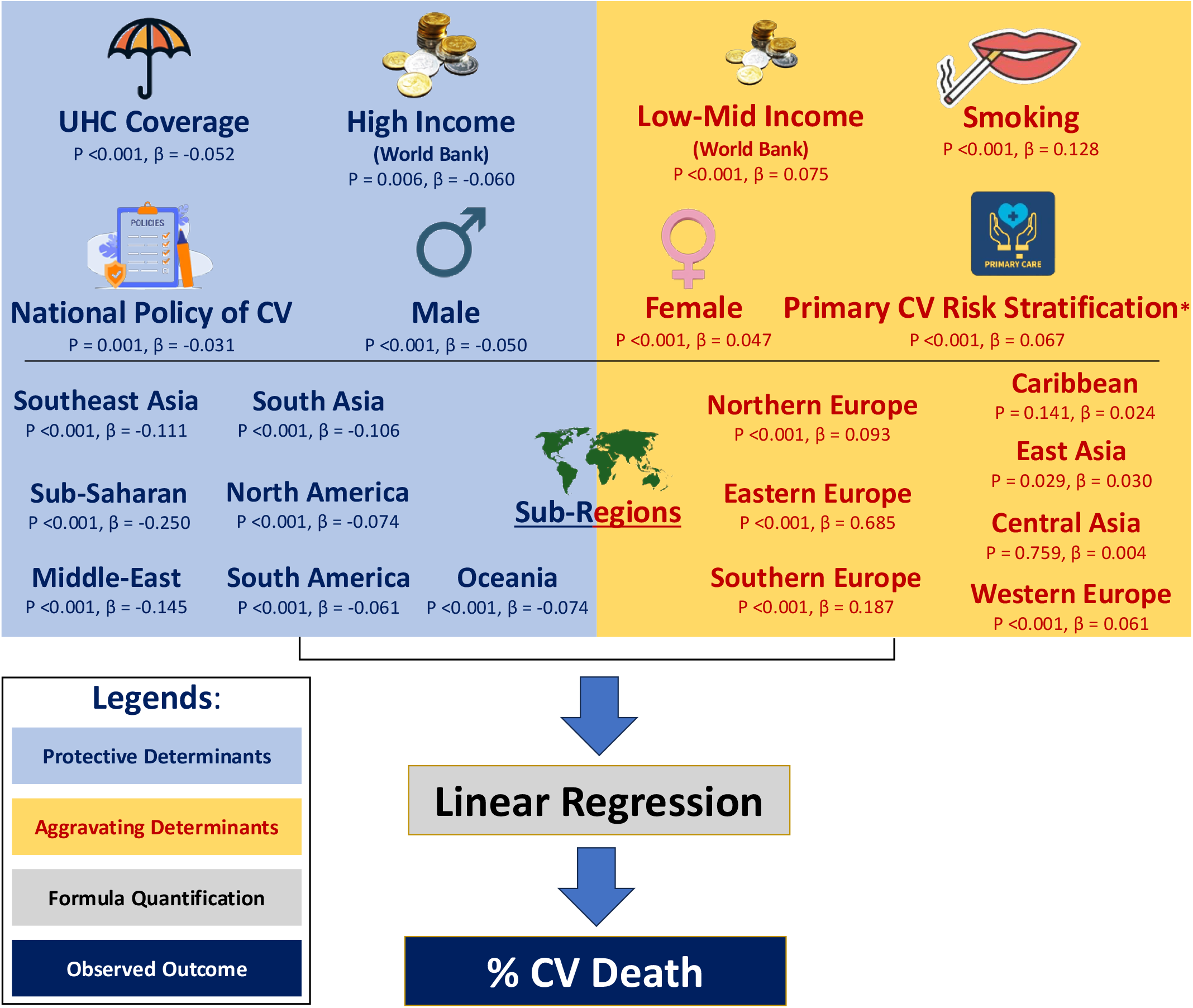

## Introduction

Cardiovascular diseases (CVDs) remain a predominant contributor to global mortality, exhibiting pronounced disparities across geographic regions and economic strata. The World Health Organization estimates that CVDs account for more than 17 million deaths annually, representing approximately 31% of worldwide mortality^1^. The nature of multivariable determinants, encompassing determinants of socioeconomic status^2^, healthcare access^3^, lifestyle behaviors^4^, and regional inequalities^5^ complicating the decision-making process in the policy level.

Despite well-established of cardiovascular determinants, the formulation of a quantitative predictive measure of cardiovascular mortality remains challenging. Traditional risk models often do not fully capture the intricate interplay among socio-demographic factors, lifestyle behaviors, and healthcare access.^2^ In this study, we addressed this limitation by constructing a novel linear regression model incorporating critical predictors such as health coverage, World Bank income classifications^3^, regional disparities, smoking prevalence^4^, and gender differences^5^. Our comprehensive approach seeks to quantify the relative contribution of each determinant, thereby refining risk stratification and enhancing predictive accuracy. The resulting formula was intended to serve as a robust tool for both clinicians and public health policymakers, facilitating targeted interventions to mitigate the burden of cardiovascular diseases.

## Methods

Data were obtained from the World Health Organization^6^ and World Bank databases^7^. Independent variables were the percentage of Universal Health Coverage (UHC) for noncommunicable diseases, World Bank income classifications (low-income [LM], upper-middle-income [UM], and high-income [H]), sub-regional classifications, gender disparity, access to cardiovascular risk screening in primary care (defined as ≥50% coverage), the presence of an operational policy/action plan for cardiovascular health, smoking prevalence, and the existence of national guidelines specified for cardiovascular disease.

The dependent variable, percentage of cardiovascular death, represents the annual proportion of cardiovascular deaths relative to the total population in each country. Baseline characteristics were summarized using medians with interquartile ranges for continuous variables and frequencies with percentages for categorical variables. Bivariate analyses were conducted using the Kruskal-Wallis test to assess differences across sub-regional classifications, and the Mann-Whitney U test for comparisons of the remaining categorical variables.

Subsequently, a linear regression model was developed using SPSS to quantify the relationships between the independent variables and cardiovascular mortality. Model diagnostics were performed to evaluate the assumptions of linearity, independence, homoscedasticity, and normality of residuals. Statistical significance was established at a two-tailed p-value of less than 0.05.

## Results

### Baseline characteristics and bivariate analyses

In the bivariate analysis, we examined the association between various determinants and the percentage of CV-related deaths (% CV death) in the study population. The median % CV death was 0.19%, with a range from 0.02% to 1.03%.

**Table 1.** CV death % amongst the included population.

| Variable | Median (min-max) in % |
| --- | --- |
| CV Death % | 0.19 (0.02-1.03) |

There was a significant relationship between income level and % CV death (P < 0.001). The proportion of included data points has been visualized in Table 2.

**Table 2.** Bivariate analysis of World Bank income classification towards % CV death.

| Determinants | n (%) | P-value |
| --- | --- | --- |
| High Income | 780 (28.4%) | <0.001 |
| Upper-Middle Income | 774 (28.2%) |  |
| Low-Mid Income | 747 (27.2%) |  |
| Low Income | 444 (16.2%) |  |

More than half of the population was covered by well-established national policies on cardiovascular (CV) health, with 63.4% receiving coverage under the National Policy of CV and 82.5% under the National CV Guidelines. Both the National Policy and National Guidelines were significantly associated with % CV death. However, CV risk stratification remains underdeveloped in many countries, with only 15.3% of the population benefiting from a comprehensive primary care approach to CV risk assessment and management. Although non-significant, female gender showed a p value of 0.25, enabling the imputation into the linear regression model.

**Table 3.** Bivariate analysis of the remaining determinants towards % CV death.

| Variable | n (%) | P-value |
| --- | --- | --- |
| National Policy of CV | 1740 (63.4%) | <0.001 |
| National Guideline | 2265 (82.5%) | <0.001 |
| Female Gender | 915 (33.3) | 0.25 |
| Primary CV risk stratification* | 421 (15.3%) | <0.001 |

\*in $\geq 50\%$ primary health care
Varied UHC and smoking prevalence were observed with a median percentage of 62% and 16.4%, respectively.

**Table 4.** Bivariate analysis of the continuous variables towards % CV death.

| Variable | Median (min-max) in % | P-value |
| --- | --- | --- |
| % UHC Coverage | 62 (16-85) | <0.001 |
| % Smoking Prevalence | 16.4 (0.1-72.8) | <0.001 |

### Linear Regression

Further analysis with linear regression was employed for variables showing a significance with a p-value of 0.25 or lesser. The linear regression was conducted to examine the relationship between various CV death determinants and the percentage of CV-related deaths in the population. The model summary reveals that the regression model has a high degree of fit, with an R value of 0.897 and an R^2^ of 0.805, indicating that the model explains 80.5% of the variance in CV death percentage. The adjusted R^2^ of 0.803 further supports the robustness of the model, accounting for the number of predictors used. The standard error of the estimate is 0.08918, reflecting the precision of the model’s predictions. The Durbin-Watson statistic of 0.428 suggests some autocorrelation in the residuals, although this does not appear to significantly affect the model’s validity.

**Table 5.**
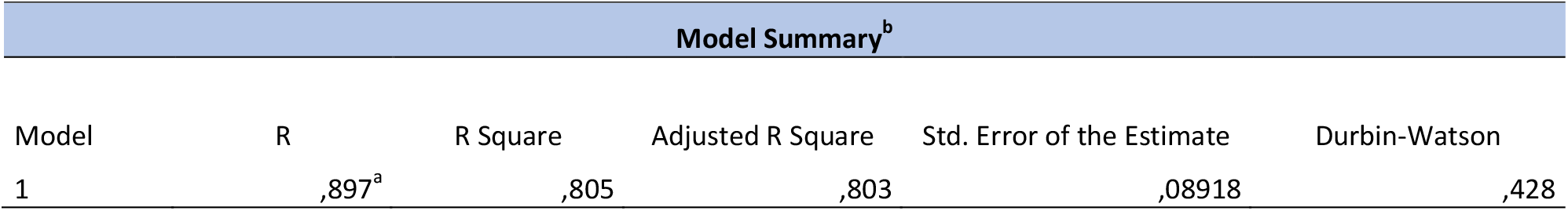
Model summary of R and R square value.

The ANOVA results show that the overall model is statistically significant, with a p-value of 0.000 (F = 404.379). The sum of squares for regression is 77,194, explaining a substantial portion of the variability in the dependent variable, while the residual sum of squares is 18,731. The large F-value and small p-value indicate that the model provides a statistically significant explanation of the variation in CV death percentages across the population.

**Table 6.**
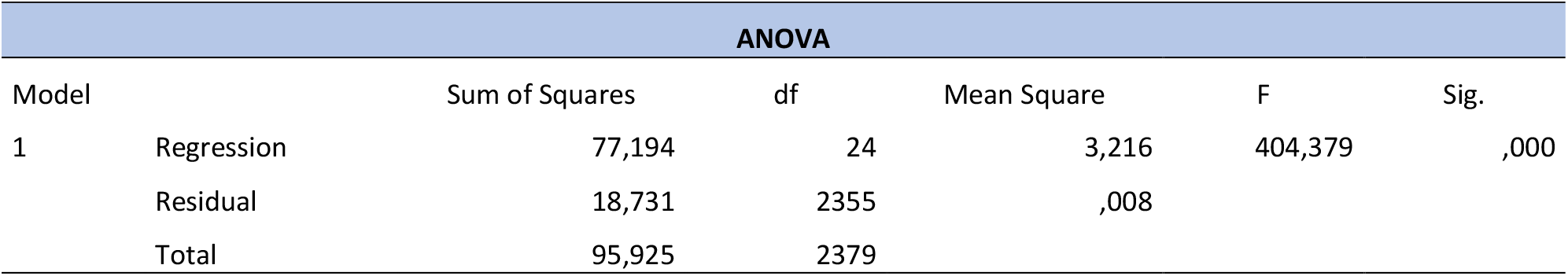
ANOVA result of regression.

The results of the linear regression analysis are presented in the coefficients table, which provides both unstandardized and standardized coefficients for the predictors of CV death percentage. The unstandardized coefficients (B) represent the change in the dependent variable for a one-unit change in the predictor, while the standardized coefficients (Beta) allow for comparison of the relative importance of each predictor.

Among the significant predictors, UHC coverage showed a negative association with CV death percentage (B = -0.001, Beta = -0.052, p < 0.001), indicating that higher UHC coverage is associated with a lower predicted CV mortality. Smoking percentage had a significant positive impact (B = 0.002, Beta = 0.128, p < 0.001), indicating a catastrophic effect of smoking in cardiovascular mortality across diverse populations. Notably, the coefficients for the regions reflect the varying impact of geographical locations on CV death percentage.

Other predictors such as gender, operational policy action plans, and the availability of cardiovascular risk stratification in primary care also contributed significantly to explaining the variance in CV death percentage. In contrast, the National Guideline was not statistically significant (p = 0.224) in predicting cardiovascular mortality.

**Figure 2.**
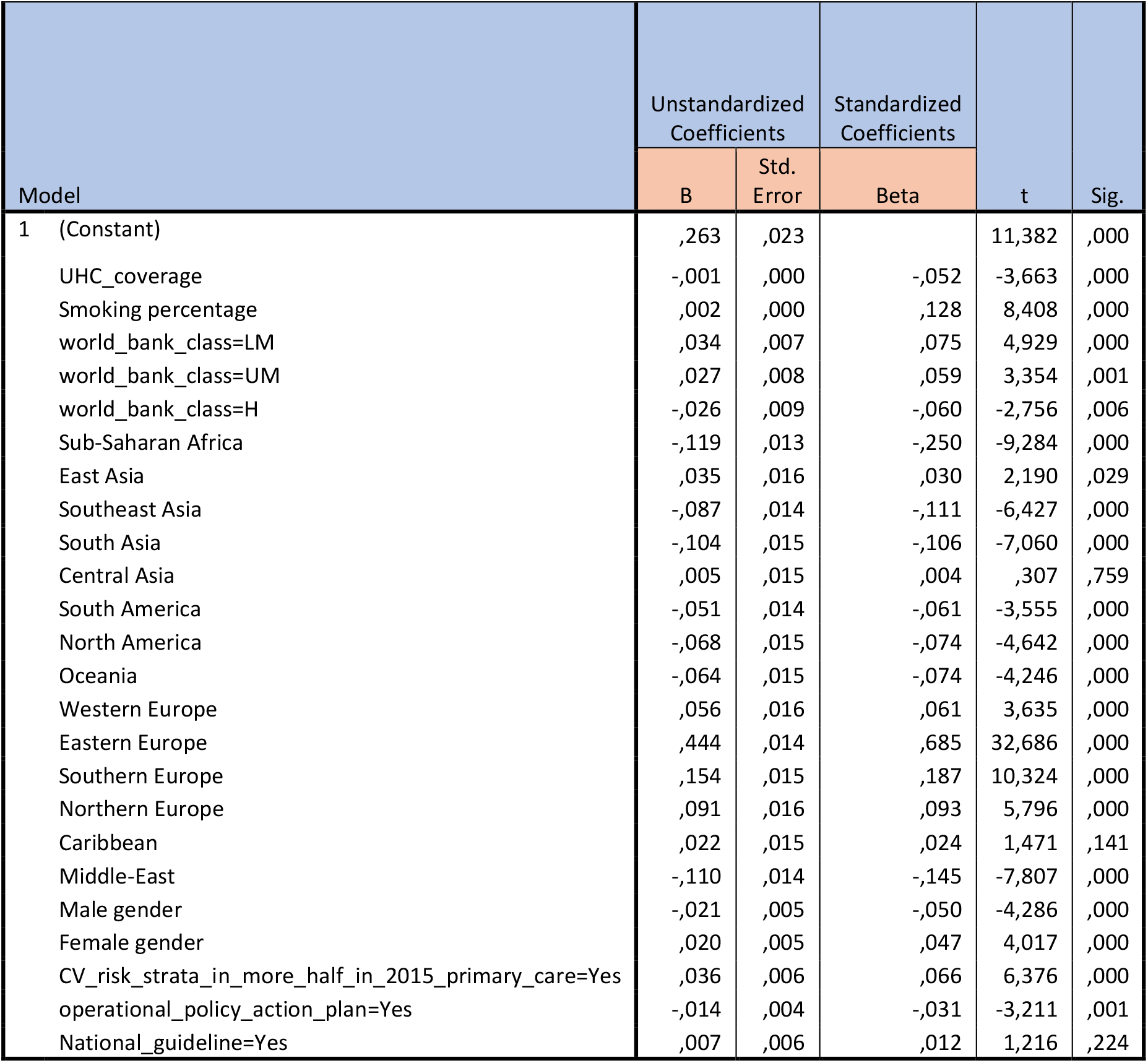
Coefficient, B, beta, t statistic and p-values result of the regression.

Residual statistics indicate the predicted values for CV death percentage, ranging from a minimum of 0.0608 to a maximum of 0.7966, with a mean predicted value of 0.2529. The residuals showed a mean of 0, with a standard deviation of 0.08873. These findings further highlight the overall robustness of the model and the importance of the determinants in predicting CV-related mortality across different regions.

**Table 7.** Residual statistics of the regression.

| Residuals Statistics<sup>a</sup> |  |  |  |  |  |
| --- | --- | --- | --- | --- | --- |
|  | Minimum | Maximum | Mean | Std. Deviation | N |
| Predicted Value | ,0608 | ,7966 | ,2529 | ,18013 | 2380 |
| Residual | -,38655 | ,34705 | ,00000 | ,08873 | 2380 |
| Std. Predicted Value | -1,067 | 3,018 | ,000 | 1,000 | 2380 |
| Std. Residual | -4,334 | 3,891 | ,000 | ,995 | 2380 |
| a. Dependent Variable: percentage_cv_death_population |  |  |  |  |  |

The final model for predicting the percentage of cardiovascular deaths in the population is as follows:

**Figure 3.**
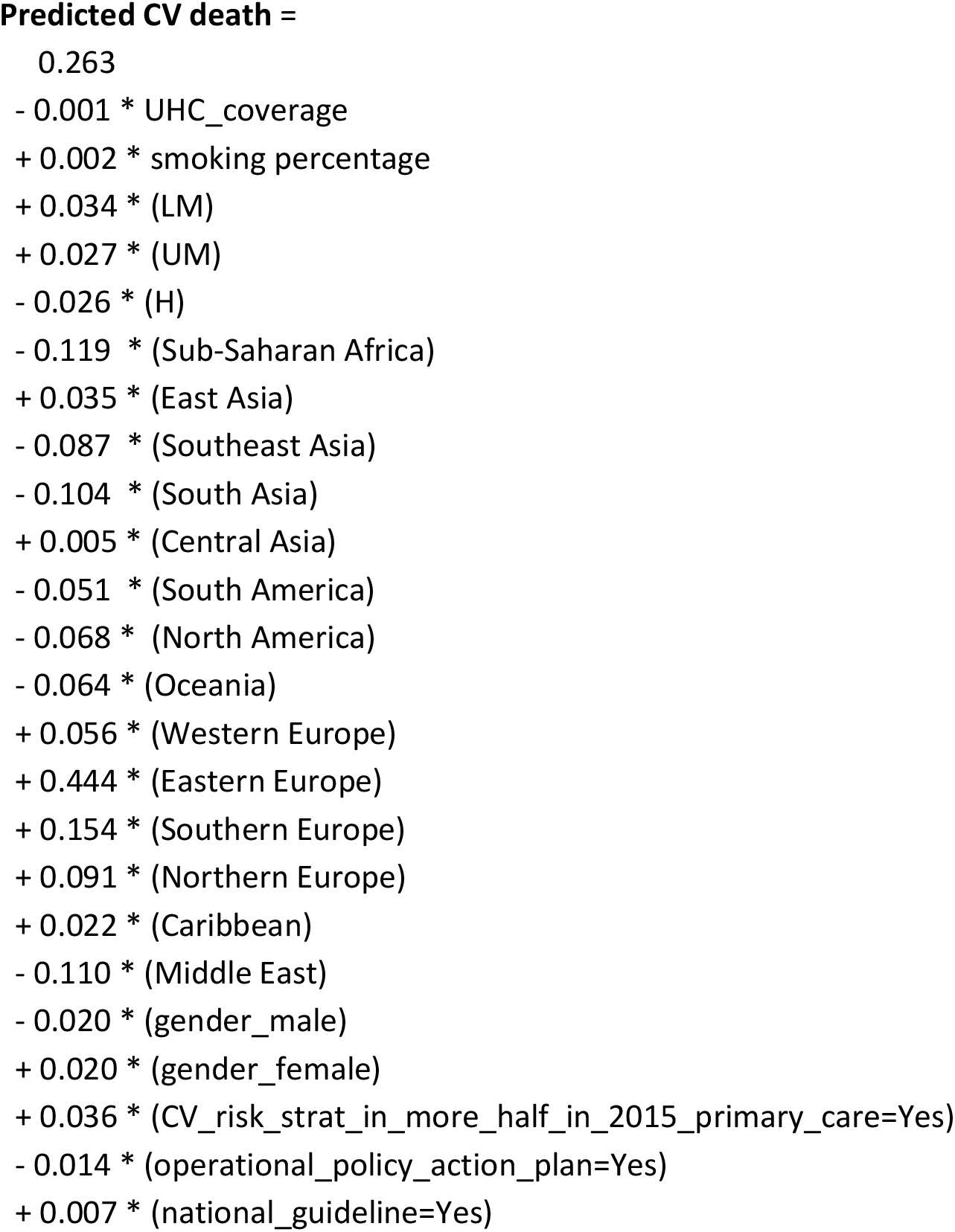
Linear regression formula to predict annual % CV death in a country with respective variables. UHC: universal health coverage; LM: lower middle income (World Bank Class); UM: upper middle income; H: high income. *Multiplication

## Discussion

This study advances current understanding by developing a novel quantitative predictive model to estimate cardiovascular (CV) mortality across 183 countries through multivariate linear regression. Our approach integrates multiple established determinants—including socioeconomic status, healthcare access, regional factors, health policies, gender, and lifestyle behaviors—into a unified, quantitative predictive formula. By quantifying the individual and collective impacts of these determinants, our model provides actionable insights to guide targeted interventions, policies, and clinical practices at the national level.

### Universal Health Coverage (UHC)

Our analysis demonstrated a significant negative association between UHC and cardiovascular mortality, consistent with previous findings highlighting the critical role of equitable healthcare access in reducing disease burden and mortality^2,8^. This underscores that countries investing in comprehensive healthcare coverage are likely to achieve improved cardiovascular outcomes, primarily through preventive services, early detection, and consistent disease management^9,10^. Policymakers at the national level should thus prioritize policies that advance universal coverage and equitable access to essential cardiovascular healthcare services.

### World Bank Income Classifications

The disparities observed across World Bank income classifications reflect the complex interplay between economic development and cardiovascular mortality. High-income nations exhibited lower cardiovascular mortality compared to low- and middle-income countries (LMICs), a finding aligning with previous studies demonstrating that economic resources are strongly linked to better health outcomes due to improved healthcare infrastructure, public health interventions, and lifestyle modifications^3,11,12^. Consistent with the findings of the Lancet NCDI Poverty Commission, our results emphasize the urgent need for context-specific strategies in LMICs that address financial constraints and healthcare inequities to mitigate the persistent disparities in cardiovascular outcomes^12^. Consequently, national governments should focus not only on healthcare investment but also on broader socioeconomic policies designed to alleviate poverty and enhance population-level cardiovascular health.

### Regional Disparities

Significant regional disparities in cardiovascular mortality rates were observed in our analysis, with higher predicted CV death percentages in regions such as Northern Europe, Eastern Europe, Southern Europe, the Caribbean, East Asia, Central Asia, and Western Europe. These regions demonstrate consistently higher cardiovascular mortality, likely reflecting the complex interplay of socioeconomic factors, healthcare access, lifestyle behaviors (e.g., diet, smoking, physical activity), and the prevalence of cardiovascular risk factors (e.g., hypertension, diabetes). Notably, Northern and Eastern Europe have been identified in previous studies as regions with alarmingly high rates of cardiovascular disease (CVD), often attributed to high smoking rates, suboptimal dietary patterns, and limited access to advanced healthcare technologies in certain areas (EuroHeart Survey, 2019)^13^. Our results align with these findings, reinforcing the need for targeted intervention strategies in these regions.

Conversely, regions such as Southeast Asia, South Asia, Sub-Saharan Africa, North America, the Middle East, South America, and Oceania exhibited lower predicted CV death rates. This finding is consistent with several large-scale studies, such as the Global Burden of Disease study, which highlights lower mortality in many parts of Southeast Asia and Sub-Saharan Africa due to relatively lower rates of non-communicable diseases (NCDs) in these regions compared to high-income countries^14^. However, it is essential to note that although these regions show lower predicted mortality, the increasing prevalence of risk factors such as smoking, poor dietary habits, and a rising burden of obesity in regions like South Asia and the Middle East may contribute to future shifts in cardiovascular mortality patterns.

An intriguing observation in our study is the low cardiovascular mortality in regions with high-risk behaviors, such as smoking, which is commonly associated with increased CVD risk. This could suggest a potential underdiagnosis or currently poor detection system for CVD-related deaths in these regions. In areas with limited healthcare infrastructure or insufficient access to diagnostic tools, CVD may go undiagnosed or misclassified, leading to lower reported mortality rates. Moreover, in some low- and middle-income countries, CVD-related deaths might be attributed to other causes, such as respiratory or infectious diseases, further obscuring the true burden of CVD mortality. This highlights the importance of improving diagnostic capabilities and establishing comprehensive mortality tracking systems to more accurately capture the true impact of cardiovascular disease, particularly in regions where risk factors are prevalent but mortality rates remain unexpectedly low.

### Gender Differences

Our study observed that males exhibiting a lower predicted cardiovascular (CV) mortality despite a higher prevalence of smoking compared to females. This counterintuitive result can be attributed to a combination of biological, diagnostic, and healthcare-related factors.

From a biological perspective, women may experience greater susceptibility to cardiovascular diseases, particularly coronary artery disease, due to hormonal influences such as estrogen, which may provide some protective effect for pre-menopausal women^15^. This could explain the higher CV mortality in females despite their lower smoking rates.

Furthermore, gender differences in healthcare access and diagnostic practices could contribute to the observed disparity. Women are more likely to present with atypical symptoms of CVD, which may lead to delays in diagnosis and treatment^16^. This delayed intervention may result in poorer outcomes for women, reflected in higher predicted CV mortality^16^. In contrast, men, who typically present with more recognizable symptoms, may receive earlier diagnosis and treatment, mitigating the impact of smoking on their CV mortality^16^.

Moreover, regional and healthcare system disparities may influence these outcomes. Men may benefit from more aggressive treatment protocols and more prompt interventions due to better recognition of their symptoms, while women may face delays in both diagnosis and care^16^.

### Risk Stratification in Primary Health Care

Interestingly, improved risk stratification program for cardiovascular disease in primary care was significantly associated with more cardiovascular mortality. This is might be due to more extensive detection of CVD in the primary healthcare, hence, better documentation on the follow-up of the mortality cases attributable to CVD. This inverse relationship substantiates that robust primary healthcare infrastructure is essential for early detection, effective management, and continuous monitoring of cardiovascular risk factors, which collectively contribute to improved health outcomes. The integration of cardiovascular screening and risk stratification within primary care settings is thus an essential component of national strategies aimed at cardiovascular risk reduction.^17^ Policymakers should prioritize investments in primary care capacity building, particularly in regions currently underserved by healthcare services, to achieve meaningful improvements in population cardiovascular health.^17^

### Operational Policy for Action Plans

Our model indicates that national policies for cardiovascular disease prevention significantly contribute to lower cardiovascular mortality rates. Such structured health policies promote consistent application of evidence-based preventive measures and facilitate systematic monitoring and evaluation of cardiovascular health outcomes^18^. National policymakers should thus prioritize the development, implementation, and enforcement of clearly articulated operational plans to provide standardized, evidence-based care across healthcare systems, ensuring more equitable health outcomes.

### Smoking Prevalence

Finally, consistent with extensive previous literatures, our study reinforced the critical role of smoking prevalence as a modifiable cardiovascular risk factor^19,20^. Countries with higher tobacco use exhibited substantially elevated cardiovascular mortality, aligning with previous global epidemiological studies demonstrating tobacco as a leading cause of preventable cardiovascular deaths^19,20^. Effective tobacco control policies remain fundamental in national strategies to reduce tobacco use and cardiovascular mortality. Policymakers should pursue aggressive and comprehensive tobacco control measures, integrating both regulatory and educational approaches, to achieve sustained cardiovascular health improvements.

### National Guideline

Our linear regression model revealed a non-significant association between the availability of national CV prevention and treatment guidelines and CV mortality rates. This finding was unexpected, given the established role of standardized guidelines in optimizing clinical outcomes for cardiovascular disease. Despite the unavailability of national CV guideline, some countries, however, may have adopted the other existing guidelines from other countries with well-established cardiovascular healthcare. Such adaptations could mitigate the expected impact of local guideline unavailability, leading to heterogeneous effects in our model.

### Strengths, Limitations, and Implications

Current study exhibits a comprehensive quantitative assessment of multiple cardiovascular determinants across diverse national settings along with predictive regression model, enabling a nuanced understanding of global disparities in the present and future. Nevertheless, limitations include the cross-sectional nature of the study, which limits causal inference, and potential data variability due to differences in reporting standards across countries.

The synthesis of new knowledge provided by this study offers actionable insights for national policymakers and health systems. Our findings suggest that future cardiovascular prevention strategies should integrate multifactorial determinants into comprehensive frameworks, prioritizing equitable healthcare access, tailored regional interventions, targeted socioeconomic improvements, gender-specific preventive strategies, strengthened primary care infrastructure, structured operational guidelines, and rigorous tobacco control policies. Adoption of these recommendations at national levels could substantially decrease global cardiovascular mortality, aligning with the broader goals of improving population health and achieving sustainable development.

## Conclusion

Present study highlights the importance of improving nationwide healthcare access, policymaking, and addressing personal modifiable lifestyle risk factors to reduce the global burden of CVD. Future research should focus on tailored, multifaceted approaches, including improving healthcare access, implementing regional interventions, targeting socioeconomic disparities, developing gender-specific prevention plans, strengthening primary care, establishing clear guidelines, and enforcing stronger tobacco control policies.

## Acknowledgement

The authors would like to thank all the research staffs in the Department of Vascular Medicine, National Cardiovascular Center Harapan Kita for their technical assistance in the data gathering and refinements.

## Funding

The authors received no funding for this research.

## Conflict of interest

All authors declare no financial or personal relationships with other people or organizations that could inappropriately influence (bias) this work. This includes no receipt of funding, grants, honoraria, stock ownership, royalties, consultancies, or any other interests related to the submitted manuscript.

## Data availability

The data may become available upon reasonable request.

## Notes

**Conflict of Interest:** None declared.

### Competing Interest Statement

The authors have declared no competing interest.

### Clinical Trial

This study does not require registration with a recognized trial registry because it is not a clinical trial.

### Funding Statement

No external funding was received for this work.

### Author Declarations

The research described in this preprint was determined to be exempt from Institutional Review Board (IRB) review because it involved the analysis of a publicly available data, and did not involve direct interaction with human subjects.

## References

[1] World Health Organization. Cardiovascular diseases (CVDs). World Health Organization website. Accessed March 23, 2025. https://www.who.int/health-topics/cardiovascular-diseases.

[2] Powell-Wiley TM, Baumer Y, Baah FO, et al. Social Determinants of Cardiovascular Disease. Circ Res. 2022;130(5):782–799. doi:10.1161/CIRCRESAHA.121.319811.

[3] World Bank. World Bank country and lending groups. World Bank website. Accessed March 23, 2025. https://datahelpdesk.worldbank.org/knowledgebase/articles/906519.

[4] World Health Organization. Tobacco. https://www.who.int/news-room/fact-sheets/detail/tobacco. Accessed March 23, 2025. (Updated 2021).

[5] Mosca L, Barrett-Connor E, Kass Wenger N. Sex/gender differences in cardiovascular disease prevention: What a difference a decade makes. Circulation. 2011;124(19):2145–2154. doi:10.1161/CIRCULATIONAHA.110.968792.

[6] World Health Organization. The Global Health Observatory. Accessed on March 23 2025. Available from: https://www.who.int/data/gho/data/indicators.

[7] World Bank. World Bank country and lending groups. Accessed March 23, 2025. Available from: https://datahelpdesk.worldbank.org/knowledgebase/articles/906519.

[8] World Health Organization. Universal Health Coverage (UHC). https://www.who.int/news-room/fact-sheets/detail/universal-health-coverage-(uhc). Accessed March 23, 2025.

[9] Starfield B. Primary care and equity in health: the importance to effectiveness and equity of responsiveness to peoples’ needs. Humanit Soc. 2009;33(1-2):56–73. doi:10.1177/016059760903300105.

[10] Kodali PB. Achieving Universal Health Coverage in Low-and Middle-Income Countries: Challenges for Policy Post-Pandemic and Beyond. Risk Manag Healthc Policy. 2023;16:607–621. doi:10.2147/RMHP.S366759.

[11] Marmot M. Social determinants of health inequalities. Lancet. 2005;365(9464):1099–1104. doi:10.1016/S0140-6736(05)71146-6.

[12] Bukhman G, Mocumbi AO, Atun R, et al. The Lancet NCDI Poverty Commission: bridging a gap in universal health coverage for the poorest billion. The Lancet. 2020;396(10256):991–1044. doi:10.1016/S0140-6736(20)31907-3

[13] Gaede L, Sitges M, Neil J, et al. European heart health survey 2019. Clin Cardiol. 2020;43(12):1539–1546. doi:10.1002/clc.23478

[14] Global burden of 369 diseases and injuries in 204 countries and territories, 1990–2019: a systematic analysis for the Global Burden of Disease Study 2019.

[15] Bairey Merz CN, Shaw LJ, Reis SE, et al. Insights from the NHLBI-sponsored Women’s Ischemia Syndrome Evaluation (WISE) study. Part II: Gender differences in presentation, diagnosis, and outcome with regard to gender-based pathophysiology of atherosclerosis and macrovascular and microvascular coronary disease. J Am Coll Cardiol. 2006;47(3 SUPPL.):pS21-S29. doi:10.1016/j.jacc.2004.12.084

[16] Elbarbary M, Shalaby HK, Elshokafy SM, Khalil MA. Gender differences in presentation, management, and outcomes among Egyptian patients with acute coronary syndrome: a single-centre registry. BMC Cardiovasc Disord. 2024;24(1). doi:10.1186/s12872-024-03996-8

[17] Yacaman Mendez D, Zhou M, Brynedal B, et al. Risk stratification for cardiovascular disease: a comparative analysis of cluster analysis and traditional prediction models. Eur J Prev Cardiol. Published online January 15, 2025. doi:10.1093/eurjpc/zwaf013

[18] Fuster Valentin, Kelly BBurke. Promoting Cardiovascular Health in the Developing World : A Critical Challenge to Achieve Global Health. National Academies Press; 2010.

[19] Zhu S, Gao J, Zhang L, et al. Global, regional, and national cardiovascular disease burden attributable to smoking from 1990 to 2021: Findings from the GBD 2021 Study. Tob Induc Dis. 2025;23. doi:10.18332/tid/200072

[20] Banks E, Joshy G, Korda RJ, et al. Tobacco smoking and risk of 36 cardiovascular disease subtypes: Fatal and non-fatal outcomes in a large prospective Australian study. BMC Med. 2019;17(1). doi:10.1186/s12916-019-1351-4

